# Initial experience in Mexico with convalescent plasma in COVID-19 patients with severe respiratory failure, a retrospective case series

**DOI:** 10.1101/2020.07.14.20144469

**Authors:** Michel F. Martinez-Resendez, Fernando Castilleja-Leal, Alejandro Torres-Quintanilla, Augusto Rojas-Martinez, Gerardo Garcia-Rivas, Rocio Ortiz-Lopez, Victor Trevino, Reynaldo Lara-Medrano, Hiram Villanueva-Lozano, Teresa Ramirez-Elizondo, Victor M. Sanchez-Nava, Francisco Moreno-Hoyos, Alfonso Martinez-Thomae, Martin Hernández-Torre, Carlos Diaz-Olachea, Servando Cardona-Huerta, Sylvia de la Rosa-Pacheco, Carlos Diaz-Garza, Paola Reynoso-Lobo, Alma R. Marroquin-Escamilla, Jessica G. Herrera-Gamboa, Fatima M. Alvarado-Monroy, Claudia D. Aguayo-Millan, Francisco F. Villegas-Macedo, Jesus Efrain Flores-Osorio, Daniel Davila-Gonzalez, María Eugenia Diaz-Sanchez, Guillermo Torre-Amione

## Abstract

**Introduction:** Hospital mortality due to COVID-19 in Mexico is high (32%) and as of today, effective treatment options are limited. More effective treatments that shorten hospital stay and reduce mortality are needed. Initial reports for the use of convalescent plasma (CP) therapy for COVID-19 appear promising. We describe a case series of eight patients with impending respiratory failure, who underwent CP therapy.

**Methods:** Six male and two female (ages 31 to 79) patients that were admitted to the intensive-care unit for severe COVID-19 were transfused with two doses of CP (250 mL per dose, anti-SARS-CoV-2 IgG titers > 1:100). Donors were six SARS-CoV-2 infected males who remained asymptomatic for > 7 days and were negative for two nasopharyngeal RT-PCR tests. Clinical characteristics, inflammatory and cellular injury markers, chest X-ray findings and viral loads were analyzed before and after CP administration. Viral load association to disease severity was further analyzed on a separate cohort of asymptomatic vs hospitalized patients with COVID-19.

**Results:** Eight patients with respiratory failure were successfully discharged with a median length of stay of 22.5 (IQR 18.25-29.00). After CP therapy, we observed a reduction of C-reactive protein (CRP) (median, 22.80 mg/dL vs. 1.63 mg/dL), and of procalcitonin (median, 0.27 ng/mL vs. 0.13 ng/mL). High-Sensitivity Cardiac Troponin I (hs-cTnI), Brain Natriuretic Peptide (BNP) and Lactate Dehydrogenase (LDH) were lower, and a mild reduction of pulmonary infiltrates by chest X-ray was observed. Lastly, a reduction of viral load was after CP therapy was found. (log, median [IQR], 1.2 [0.70-2.20] vs. 0.25 [0.00-1.78]). We observed no adverse effects.

**Conclusions:** CP could potentially be an effective therapeutic option for patients with severe COVID-19. Clinical benefit needs to be studied further through randomized controlled trials.

## Introduction

As the COVID-19 pandemic has progressed, it has shifted epicenter into the Americas. As of June 9, 4 of the 10 nations with the highest number of new cases are Latin American countries: Brazil, Peru, Chile and Mexico (1). Complicating things further, Mexico has seen an increasing hospital mortality, which stands now at 32% (2). Hospital saturation must be an involved factor explaining said high mortality, as Mexico reports the lowest number of hospital beds per 1,000 habitants among the OECD member countries, as per the latest data (3). Certainly, therapies that reduce mortality and shorten hospital stay are needed globally. As there are few therapeutic options, such as antivirals and monoclonal antibodies, and their potential efficacy is still being determined, convalescent plasma (CP) could represent a lower cost alternative (4). This last property could be particularly helpful for developing countries, if CP is proven to be effective.

As CP has been used for more than a century for the treatment of different infectious diseases, especially during epidemics or pandemics such as SARS, influenza or Ebola virus, there is vast experience with this safe therapeutic alternative. Nonetheless, the mechanisms of action may vary for every virus for which it is used, and are still being revisited (5). CP has been successfully used in the treatment of SARS and MERS. Moreover, recent independent studies of CP therapy in patients with COVID-19 provide evidence for its use (6–8). In this paper we describe our initial experience with the use of CP in a case series of patients with COVID-19 with impending respiratory failure.

## Materials and methods

### Patients

Eight critically ill patients with COVID-19 that were admitted to the COVID-19 Unit at Hospital San José TecSalud in Monterrey, Mexico, from March 10 to May 21, 2020, received CP therapy. All patients had impending respiratory failure associated with COVID-19 and were confirmed by RT-PCR testing before received treatment with CP. ABO and Rh blood types of the patients were determined for potential compatibility with the CP donor, and each subject received two transfusions of 250 mL of CP separated by 24 hours. Plasma was infused at 20 mL/min and monitored for 15 minutes after the infusion and no premedication was used. All patients were previously initiated on local established and protocolized therapy including antiviral agents, moreover the patients received supportive care such as antibiotic treatment, antipyretics, analgesics, fluids and nutrition. This study was approved by the Clinical Research Ethics Committee of the School of Medicine at Tecnológico de Monterrey (P000353-COVID-19 TecSalud-CEIC-CR001). Written informed consent was obtained from all patients. All patients received CP treatment under compassionate use due to impending respiratory failure after case review by the hospital bioethics committee.

### Donors

We obtained CP from 6 male donors who were between the ages of 22 and 57 years. The donors had recovered from SARS-CoV-2 infection with at least 7 days without any symptoms and 2 negative RT-PCR tests. They were invited to donate their plasma after written informed consent was obtained. In addition, the donors were screened to rule out hepatitis B virus, hepatitis C virus, Epstein Barr virus, HIV, *Brucella abortus, Trypanosoma cruzi* and syphilis infection at the time of blood donation. Donors showed anti-SARS-CoV-2 IgG antibody titers higher than 1:100 as determined by ELISA. Written informed consent was obtained and ABO-compatible plasma samples were obtained by apheresis. Depending on bodyweight, 600-800 mL were collected and divided into 250 mL aliquots. A first dose of CP was transfused to recipients at baseline and a second dose 24 hours afterwards.

### Clinical information

Data from patients was obtained from medical records. Retrieved data included: demographics, symptoms, days from symptom onset to admission, length of stay, pharmacologic therapy, type and duration of ventilator support, vital signs (temperature, heart rate, respiratory rate, SaO_2_/FiO_2_, and blood pressure), severity scores [Sequential Organ Failure Assessment (SOFA), the quick Sequential Organ Failure Assessment (qSOFA), the Pneumonia Severity Index (PSI), the Confusion, blood Urea nitrogen, Respiratory rate, Blood pressure, age 65 score (CURB-65)], laboratory data [complete blood count, blood chemistry, liver function testing, cultures, SARS-CoV-2 transcript copies, inflammatory markers like C-reactive protein (CRP), procalcitonin, troponin, brain natriuretic peptide (BNP), lactate dehydrogenase (LDH), D-dimer, ferritin and interleukin-6 (IL-6)], chest X-Ray or CT-scan findings, and clinical events and outcomes (ARDS, bacterial pneumonia, multiple organ dysfunction syndrome and death). Inflammation markers were taken at admission, before each dose of CP and at 48 hours follow-up.

### Patient cohort for viral load control group

Discarded samples from clinical laboratory were obtained to determine viral load in a group of asymptomatic outpatients (n=23) as well as in hospitalized COVID-19 patients. (n=21).

### Chest X-ray pulmonary infiltrates severity scoring

All patients were evaluated for pulmonary infiltrates severity on chest X-ray as previously reported (9). Briefly, each lung is assigned a score of 0-4 depending on the extent of consolidation or ground-glass opacities as follows: 0 = no involvement; 1 = <25%; 2 = 25-50%; 3 = 50-75%; 4 = >75% involvement. The final score corresponds to the sum of the scores for each lung.

### RT-PCR Test

We used the Applied Biosystems™ TaqMan™ 2019-nCoV Assay Kit v1 (Cat. No. A47532) for the qualitative detection and characterization of 2019-nCoV RNA. The kit includes three PCR assays that target viral genes, and one positive control assay that targets the Human RNase P RPPH1 gene. The reactions were performed according to manufacturer instructions. Briefly, the 25uL reactions for each 2019-nCoV assay include Master Mix 6.25uL, 2019-nCoV assay primers and probes per gene 1.25 uL, RNAse P 1.25uL, RT-PCR grade water 11.25uL, and testing sample 5uL. The testing sample could be a biological specimen from a patient, positive control (A47533), or water as non-template control. The cycling conditions were 2 min @ 25°C, 10 min @ 50°C, 2 min @ 95°C, and 40-45 cycles of 3 sec @ 95°C and 30 sec @ 60°C. This PCR program was run in a QuantStudio 5 Real-Time PCR system (ThermoFisher). Automatic threshold detection was used as instructed. A sample is positive if at least two of the three COVID-19 genes amplifications were detected above threshold before cycle 38. A negative result is given if no amplification of the three COVID-19 genes was observed. An invalid result is given if any of the three amplifications of RNAse P was not observed before cycle 38. If otherwise, an indeterminate result is given. If an indeterminate result is observed, the set of reactions for the sample is repeated and the result is reported. To estimate the viral load, we used a dilution series of the positive control (A47533) included in each PCR plate and used the regression curve to estimate the minimum and maximum viral load from the curves of each of the three viral genes. The regression was estimated automatically by the Design & Analysis Software Release Version 2.3.0 obtained from Thermo Fisher Scientific web site.

### Serology Test

Antibody titers for IgG and IgM antibodies against SARS-CoV-2 were obtained using the EDI™ Novel Coronavirus COVID-19 IgG and IgM ELISA Kits (Epitope Diagnostics, San Diego, CA) following manufacturer instructions. Briefly, serum samples were diluted (1:50 and 1:100) and incubated in the microtiter wells of a microplate that is coated with COVID recombinant full length nucleocapsid protein (SARS-CoV-2 antigen). Positive and negative control calibrators were included. Results were reported as positive or negative at the specific dilution (1:50 or 1:100 for IgG and 1:10 for IgM) based on the optical density (OD). The cutoffs were calculated considering the average value of the absorbance of the negative control (xNC). The cutoffs for IgG antibodies were calculated using the following formulas: Positive cutoff = 1.1 X (xNC + 0.18) and negative cutoff = 0.9 × (xNC + 0.18). Cutoff values < 0.284 were considered negative, > 0.284 to < 0.3476 were considered borderline, and > 0.3476 were considered positive, per manufacturer instructions. The cutoffs for IgM antibodies were calculated using the following formulas: Positive cutoff = 1.1 X (xNC + 0.10) and negative cutoff = 0.9 × (xNC + 0.10). Cutoff values < 0.218 were considered negative, > 0.218 to < 0.265 were considered borderline, and > 0.265 were considered positive, per manufacturer instructions.

### Statistical Analysis

Frequencies and percentages are used for categorical data, and medians and interquartile ranges for continuous variables. GraphPad Prism 7.0 was used for plotting graphs and statistical analysis.

## Results

### Patient characteristics

We report eight patients (2 female, 6 male) critically ill with a confirmed diagnosis of COVID-19 that were admitted to our institution from March 10, 2020 to May 21, 2020 and received CP therapy. The median age was 57 (IQR, 48-69), the median time from onset of symptoms to admission was 6 days (IQR, 3.00-8.75) and the median length of stay 22.5 days (IQR, 18.25-29.00) (**Table 1**). The most common comorbidities present were overweight/obesity (87.5%), hypertension (50%), diabetes mellitus (25%) and smoking (12.5%). The most common symptoms on admission were fever (100%) and dyspnea (100%), followed by cough (87.5%) and myalgia/arthralgia (75%). Seven out of eight patients (87.5%) received chloroquine/hydroxychloroquine, lopinavir/ritonavir, and azithromycin. Six patients (75%) received ceftaroline and 3 patients (37.5%) tocilizumab.

**Table 1.** Baseline clinical and demographic characteristics.

| **Variable** | ***n* = 8** |
| --- | --- |
| **Age, median (IQR), yr** | 57 (48-69) |
| **> 60, No. (%)** | 3 (37.5) |
| **Female, No. (%)** | 2 (25) |
| **Male, No. (%)** | 6 (75) |
| **Time from onset to admission, median (IQR), days** | 6 (3-8.75) |
| **Length of stay, median (IQR), days** | 22.5 (18.25-29) |
| **Comorbidities, No. (%)** |  |
| **Smoking** | 1 (12.5) |
| **Hypertension** | 4 (50) |
| **Diabetes** | 2 (25) |
| **Overweight/obesity** | 7 (87.5) |
| **Symptoms at admission, No. (%)** |  |
| **Fever** | 8 (100) |
| **Nasal congestion** | 1 (12.5) |
| **Headache** | 2 (25) |
| **Cough** | 7 (87.5) |
| **Sputum** | 2 (25) |
| **Odynophagia** | 3 (37.5) |
| **Dyspnea** | 8 (100) |
| **Myalgia/arthralgia** | 6 (75) |
| **Chills/diaphoresis** | 4 (50) |
| **Clinical parameters, median (IQR)** |  |
| **Heart rate, bpm** | 100 (78.75-135) |
| **Respiratory rate, rpm** | 25 (17.75-31.5) |
| **Temperature, °C** | 37.85 (36.8-38.22) |
| **SOFA, pts** | 2 (2-4.25) |
| **QSOFA, pts** | 0.5 (0-1) |
| **CURB65, pts** | 1 (0.25-1.75) |
| **PSI, pts** | 78.5 (61.5-85.75) |
| **Treatment, No. (%)** | 7 (87.5) |
| **Chloroquine/Hydroxychloroquine** | 7 (87.5) |
| **Lopinavir/Ritonavir** | 7 (87.5) |
| **Azithromycin** | 6 (75) |
| **Ceftaroline** | 3 (37.5) |

Five patients (62.5%) were under mechanical ventilation and three patients (37.5%) high-flow oxygen therapy (**Table 2**). The median duration of ventilatory/oxygen therapy was 12.5 days (IQR, 7.50-14.75). Under ventilatory/oxygen support, the median SaO_2_ was 90% (IQR, 82.50-94.25), the median FiO_2_ was 40% (IQR, 36.25-47.50) and the median SaO_2_/FiO_2_ ratio was 2.25 (IQR, 1.67-2.61). No adverse effects were observed.

**Table 2.** Baseline laboratory and respiratory parameters.

| Laboratory parameters,<br>median (IQR) | *n* = 8 |
| --- | --- |
| **Hemoglobin, g/dL** | 13.85 (12.7-14.75) |
| **Platelets, x 10<sup>3</sup> /μL** | 204.5 (193-234) |
| **Glucose, mg/dL** | 121 (86.5-142.75) |
| **AST, U/L** | 40.5 (23.5-45) |
| **ALT, U/L** | 45 (25.25-61.5) |
| **Total bilirubin, mg/dL** | 0.60 (0.3-0.9) |
| **Creatinine, mg/dL** | 0.9 (0.73-1.08) |
| **BUN, mg/dL** | 12.35 (8.75-16.45) |
| **Sodium, mEq/L** | 136.5 (134.25-140.5) |
| **Potassium, mEq/L** | 4.25 (3.8-4.65) |
| **Chloride, mEq/L** | 101.5 (98-107.07) |
| **Respiratory parameters** |  |
| **Mechanical ventilation, No. (%)** | 5 (62.5) |
| **High flow oxygen therapy, No. (%)** | 3 (37.5) |
| **Duration of ventilatory/oxygen therapy, median (IQR), days** | 12.5 (7.50-14.75) |
| **SaO<sub>2</sub>, median (IQR), %** | 90 (82.5-94.25) |
| **FiO<sub>2</sub>, median (IQR), %** | 40 (36.25-47.5) |
| **SAFI, median (IQR)** | 2.25 (1.67-2.61) |

### Laboratory parameters

Seven out of eight patients (87.5%) showed lymphocytopenia on admission and showed a slight improvement after CP therapy (median, 0.75 ⨯ 10^3^ /µL vs. 0.83 ⨯ 10^3^ /µL) (**Fig 1**). Three patients (37.5%) were found with leukocytosis on admission, and the leukocyte count increased after CP therapy (median, 7.40 ⨯ 10^3^ /µL vs. 11.70 ⨯ 10^3^ /µL). Other laboratory parameters used as markers of inflammation were found with mixed results. We observed a reduction of C-reactive protein (CRP) (median, 22.80 mg/dL vs. 1.63 mg/dL), and of procalcitonin (median, 0.27 ng/mL vs. 0.13 ng/mL) after CP therapy. On the other hand, an increase of IL-6 (median, 16.00 pg/mL vs. 65.79 pg/mL) and ferritin (median, 421.0 ng/mL vs. 919.0 ng/mL) was observed after CP therapy.

**Fig 1.**
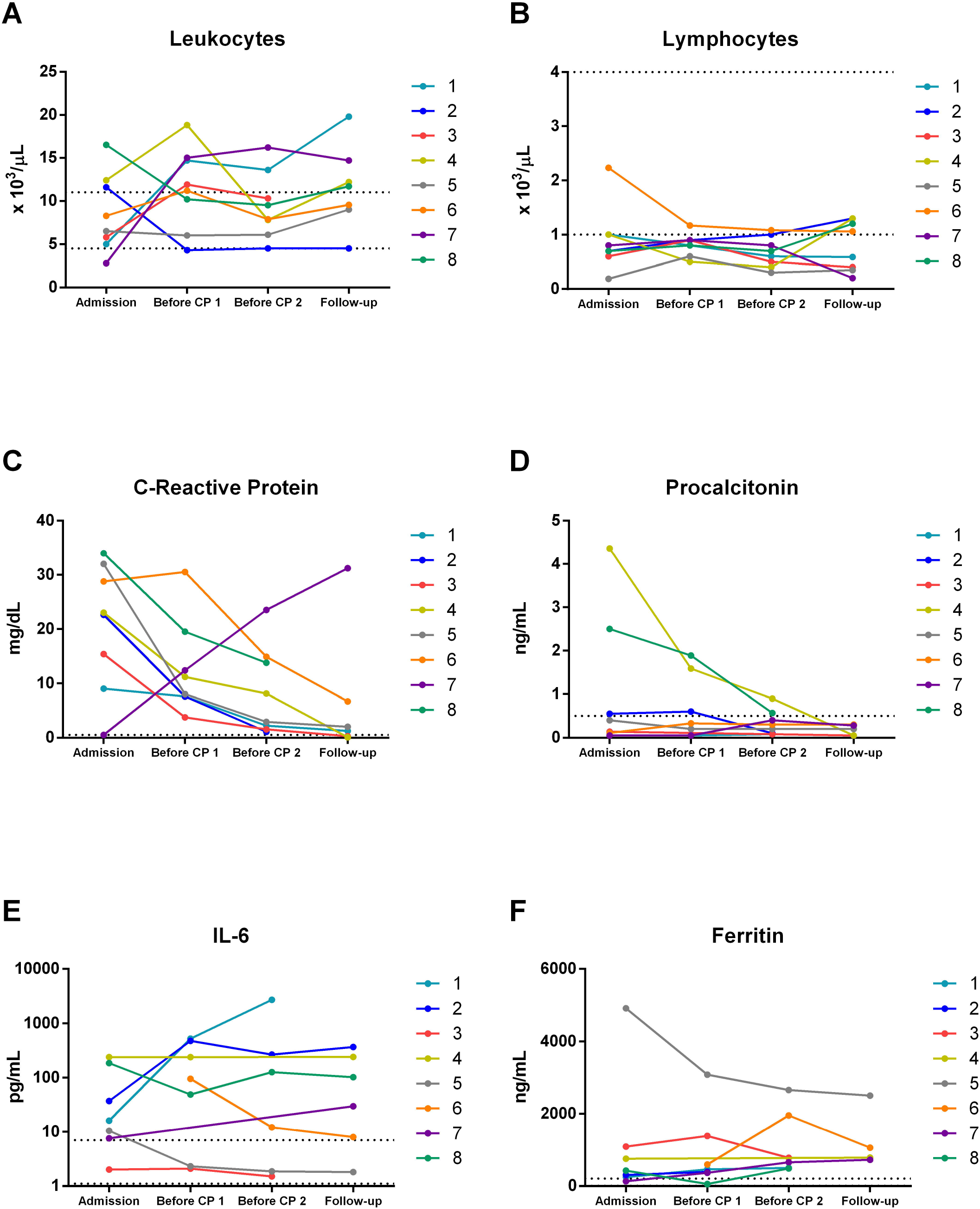
Markers of inflammation before and after CP therapy. Dynamic changes of inflammation markers before and after CP therapy in all patients. Dotted horizontal lines represent reference value ranges.

A reduction of injury markers was observed (**Fig 2**): High-Sensitivity Cardiac Troponin I (hs-cTnI) (median, 8.20 ng/L vs. 1.50 ng/L), Brain Natriuretic Peptide (BNP) (median, 50.50 pg/mL vs. 14.20 pg/mL) and Lactate Dehydrogenase (median, 368.0 U/L vs. 325.0 U/L) were lower after CP therapy. An increase of D-dimer was observed after CP therapy (median, 393 ng/mL vs. 900 ng/mL).

**Fig 2.**
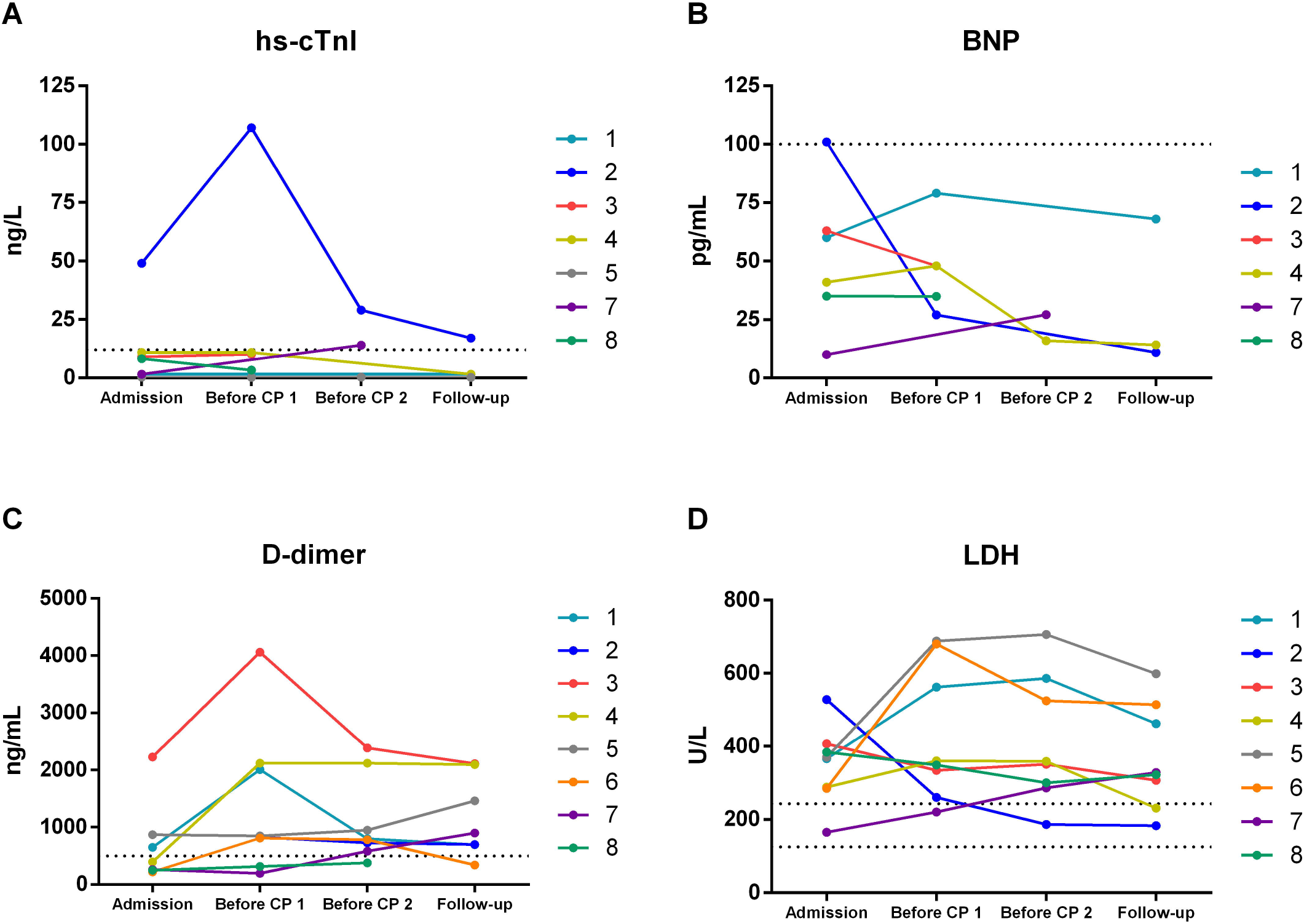
Markers of cellular injury before and after CP therapy. Dynamic changes of cellular injury markers before and after CP therapy in all patients. Dotted horizontal lines represent reference value ranges.

### Pulmonary infiltrates

A mild reduction of pulmonary infiltrates severity score by chest X-ray was observed (median, 6 vs 5). Representative chest x-ray images for two patients, along with the severity score, before and after CP therapy, are shown (**Fig 3**).

**Fig 3.**
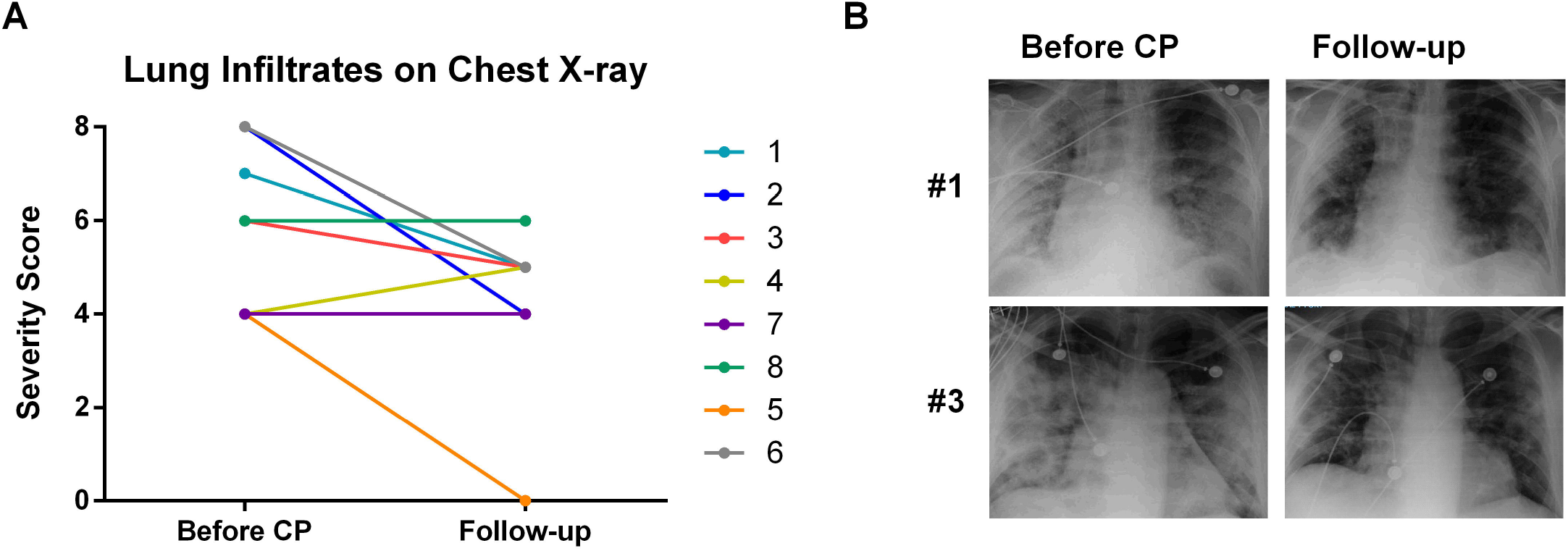
Severity of chest x-ray pulmonary infiltrates before and after CP therapy. Dynamic changes of lung infiltrates on chest X-ray before and after CP therapy. **A)** Changes in lung infiltrates as assessed by a severity score, in all patients. **B)** Representative images of the chest X-ray of two patients before and after CP therapy.

### Viral load

On a separate group of 44 patients (23 with mild/asymptomatic course, 21 with severe course) we observed an association between viral load and disease severity (**Fig 4A)**. Patients with severe COVID-19, defined as requiring hospitalization, showed a higher viral load (log, median [IQR], 0.90 [0.30-1.34] vs. 1.67 [0.74-2.22], p < 0.05). This led us to question whether viral load could be associated with clinical course after CP therapy on COVID-19 patients with severe illness. A reduction of viral load was observed in the group of eight hospitalized patients with severe COVID-19 after CP therapy (log, median [IQR], 1.2 [0.70-2.20] vs. 0.25 [0.00-1.78]) (**Fig 4B**).

**Fig 4.**
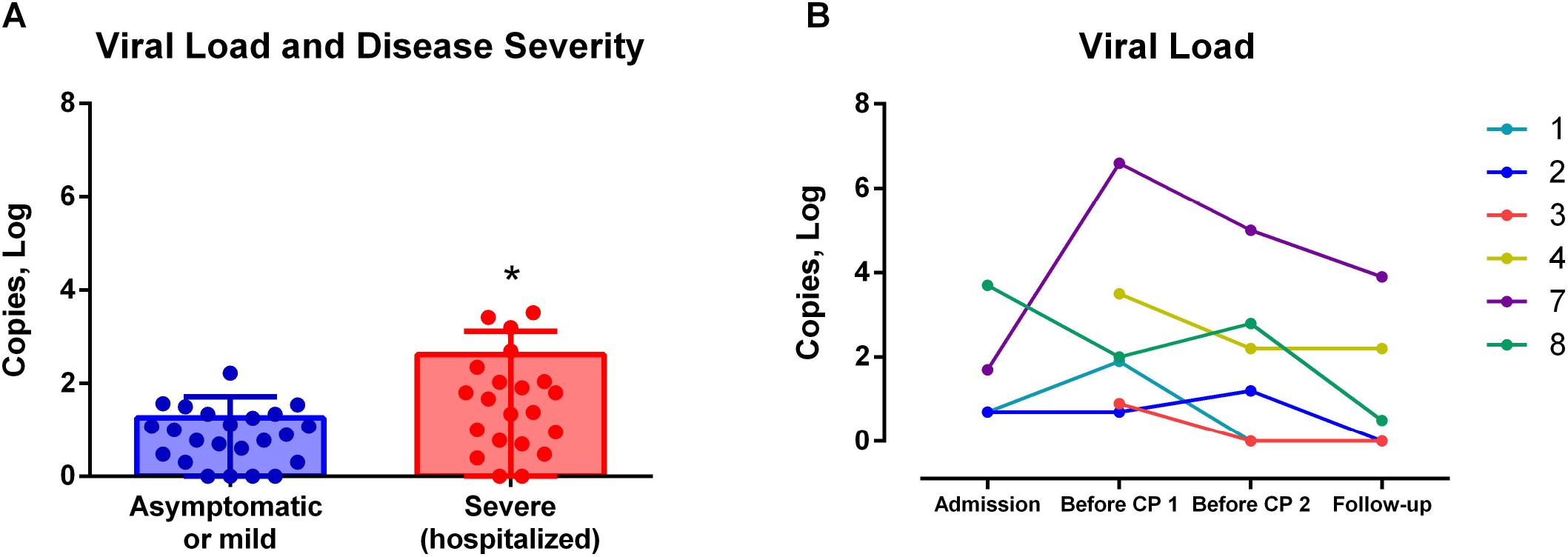
Viral load and COVID-19 disease severity. **A)** Viral load in asymptomatic/mild cases vs. hospitalized patients with severe illness, in a different study group. **B)** Dynamic changes of viral load determined before and after CP therapy for all the original group patients.

## Discussion

The major findings of this paper are: first, it was safe to administer CP to COVID-19 patients with impending respiratory failure. Second, inflammatory and cellular injury markers showed a decreasing tendency following CP administration. Third, significant improvements were observed in pulmonary infiltrates. Fourth, sequential analysis of viral load in respiratory samples showed a progressive reduction over-time. And finally, all patients were discharged home. Taken together, these observations support the hypothesis of the effectiveness of CP for COVID-19 patients with impending respiratory failure. To our knowledge, this is the first reported experience in Mexico of CP use for patients with COVID-19 with impending respiratory failure.

Experience with CP therapy is vast, as it has been utilized since the early twentieth century. During the 1918 influenza virus pandemic, CP therapy was implemented and evidence suggests it had an impact reducing mortality (10,11). Moreover, the use of CP has been evaluated for the treatment of several viral illnesses, such as Influenza A (H1N1), SARS, MERS and Ebola (12–16), and has been associated to reductions in mortality. For newly emerging viral mediated diseases, such as COVID-19, with no objective treatments, it provides at least a reasonable and testable approach. Moreover, it has been considered a lower cost alternative (4), and its use could be advantageous for developing countries, provided its efficacy is demonstrated by randomized controlled trials.

The primary hypothesis of the mechanism of action of CP therapy for COVID-19 focuses on the fact that recovered individuals have developed antibodies that can neutralize viral activity and therefore, accelerate the recovery process (5,10,17–19). But this topic has been revisited recently (5). The mechanisms of action of CP are, probably, more extensive and therefore, best described as those that may result from direct antiviral activity and those that result from immunomodulation (5).

The antiviral activity of CP, in general, is likely the result of the presence of neutralizing antibodies (NAbs) (10,18,20,21), and to our knowledge this is yet to be confirmed for COVID-19 (6). It is possible that the number of neutralizing antibodies produced during the infection, or administered with therapeutic intent, determines to some extent the disease severity and clinical outcomes.

But NAbs represent only a subset of all antibodies produced against the virus. The immunomodulatory effects of CP may result, in part, from antibodies that block the effect of autoantibodies, proinflammatory cytokines and complement activation (5). In the context of COVID-19, possible immunomodulatory effects of CP could be equally important. In this sense, COVD-19 infection is associated with a profound inflammatory response beyond the injury that results from viral cellular damage (22–27).

Various strategies directed at modulating the immune response are currently being used off-label or are being studied in clinical trials, but evidence in favor or against their use for COVID-19 is still insufficient (28). The most representative therapies that target immunomodulation are interleukin-1 (IL-1) and interleukin-6 (IL-6) inhibitors, others include interferons alfa and beta, and Janus Kinase (JAK) inhibitors. For example, Tocizilumab, an IL-6 receptor antagonist already FDA approved for rheumatoid arthritis among other indications, has shown promising results for the treatment of COVID-19 in preliminary reports (29–31).

In a recent, multicenter, randomized clinical trial, CP therapy for patients with severe COVID-19 was associated with antiviral activity, shown as a higher rate of negative SARS-CoV-2 PCR up to 72 hours later (6). The findings of our paper are consistent with this observation (**Figure 4B)**. To understand this further, we compared viral loads among asymptomatic SARS-CoV-2 PCR positive patients vs. COVID-19 hospitalized patients (**Figure 4A)**. We found a significant difference among said groups. Hospitalized patients had higher viral loads than asymptomatic ambulatory patients. This observation suggests, that at least at some point in the disease process, viral loads are associated with disease severity (32). Moreover, we observed a decreasing tendency for inflammatory markers and pulmonary infiltrates severity after CP therapy, consistent with previous reports (8).

There are limitations to our present study. Patients were receiving standard care, which could interact with the course of illness or with CP therapy. Neutralizing antibodies were not determined. However, our preliminary results support the completion of randomized larger trials.

In summary, the present study supports the observations of others and reaffirms the importance of conducting prospective clinical studies aimed to define not only the best timing and cohort of patients in which CP should be used, but also to improve our understanding of the mechanisms of action.

## Data Availability

The dataset supporting results presented here has been deposited in the Harvard Dataverse repository.

https://doi.org/10.7910/DVN/QJPC2X

## Abbreviations

COVID-19: Coronavirus Disease 2019
BNP: Brain Natriuretic Peptide
CP: Convalescent Plasma
hs-cTnI: High-sensitivity Cardiac Troponin I
IL-1: Interleukin-1
IL-6: Interleukin-6
JAK: Janus Kinase
LDH: Lactate Dehydrogenase
NAbs: Neutralizing antibodies
RT-PCR: Reverse transcription polymerase chain reaction
SARS-CoV-2: Severe acute respiratory syndrome coronavirus 2

